# COVIDFast™: A high-throughput and RNA extraction-free method for SARS-CoV-2 detection in swab (SwabFAST™) or saliva (SalivaFAST™)

**DOI:** 10.1101/2021.12.29.21268527

**Authors:** Yue Qiu, Ling Lu, Amanda Halven, Rachel Terrio, Sydney Yuldelson, Natalie Dougal, Filippo Galbo, Andrew Lu, Dexiang Gao, Bob Blomquist, Jose P. Zevallos, Brian L. Harry, Xin Yao, Shi-Long Lu

## Abstract

There is an urgent need of having a rapid, high throughput, yet accurate SARS-COV-2 PCR testing to control the COVID19 pandemic. However, the RNA extraction step in conventional PCR creates a major bottle neck in the diagnostic process. In this paper we modified the CDC COVID-19 assay and developed an RNA-extraction free RT-qPCR assay for SARS-CoV-2, i.e. COVIDFast^™^. Depending on sample types, the assay is further divided into SwabFAST^™^, which uses anterior nares nasal swab, and SalivaFAST^™^, which uses saliva. By utilizing the proprietary buffer for either swab or saliva samples, the performance of SwabFAST or SalivaFAST is equivalent to RNA-extraction SARS-CoV-2 RT-qPCR in both contrived and clinical samples. The limit of detection of either assay is 4 copies/μL. We further developed a semi-automatic system, which is easy to adapt by clinical lab for implementation of a high-throughput SARS-CoV-2 test. Working together with the COVIDCheck Colorado, we have tested over 400,000 samples using COVIDFast (83.62% SwabFAST and 16.38% SalivaFAST) in less than a year, resulting in significant clinical contribution in the battle against COVID-19 during the pandemic.

## INTRODUCTION

The rapid global spread of contagious diseases presents a major healthcare challenge. For example, the rapid spread of the severe acute respiratory syndrome coronavirus-2 (SARSCoV-2), resulting in a global pandemic, has placed an emphasis on the criticality of rapid and high-throughput detection. (*1-3*) Typically, with most respiratory viruses, people are thought to be most contagious when they are most symptomatic. With SARS-CoV-2, however, there have been reports of asymptomatic spread from infected individuals. Accordingly, to monitor the presence of SARS-CoV-2 and to prevent its spread, it is crucial to detect infection rapidly and high throughput. (*4-6*)

Current detection techniques for many infectious diseases involve the use of polymerase chain reaction (PCR). For example, various real-time PCR assays (also referred to as quantitative PCR or qPCR) for detecting SARS-CoV-2 RNA have been developed worldwide, with different targeted viral genes or regions and different sample types (*7-10*). While current PCR methods allow for the detection and diagnosis of infectious diseases, those methods suffer from drawbacks. One notable drawback is that current approaches rely on an initial step of isolating and purifying nucleic acids from a clinical sample as part of the viral testing protocol. For example, the application of qPCR for the relative quantification of an RNA typically requires: 1). the isolation and purification of total RNA from the sample; 2). elution and possible concentration of the material; and 3). the use of purified RNA in a reverse-transcription (RT) reaction resulting in complementary DNA (cDNA), which is then utilized for the qPCR reaction. The initial nucleic acid isolation and purification step (i.e., extraction step) required in conventional methods, prior to undergoing PCR, constitutes a major bottleneck in the diagnostic process (*2*), as it remains both manually laborious and expensive, and further increases the chances of accidental contamination and human error. Furthermore, in a period of high demand, a shortage of nucleic acid extraction supplies can exacerbate the limitations of such viral detection methods (*2, 11*).

We thus developed an assay namely “ COVIDFast” for rapid, high-throughput, RNA extraction-free qPCR testing of biological samples for SARS-CoV-2 detection. This assay consists of: 1). an innovative, proprietary buffer for sample transport and preparation, 2). methods for rapid, extraction-free detection and analysis of nucleic acid in a biological sample, and 3). a unique combination of sample collection kits that make high throughput sample processing and testing possible, all of which in combination overcome the shortcomings of the existing RT-qPCR detection methods. In this paper, we describe the assay “ COVIDFast” on the detection of SARS-CoV-2 in either an anterior nares swab sample (SwabFAST ^™^) or a saliva sample (SalivaFAST^™^).

## RESULTS

### High Throughput Enabled by Collection Kit and Assay Innovation

COVIDFast assay was created as a laboratory-developed-test (LDT). Therefore, there is flexibility in collection device and vessel choices, and the testing procedure setup. A notable feature of COVIDFast is its incorporation of a particular set of sample collection devices, collection vessel, and corresponding laboratory equipment that allows for automated, high-throughput testing processes and efficient use of labware (Figure 1). To this point, we uniformed a 2 mL, bar-coded, cryogenic vial (or “ cryovial”) with an externally threaded screw cap as the sample collection vessel for both SwabFAST and SalivaFAST, Using the same cryovial for both saliva and swab specimen collection facilitates downstream sample accessioning and automation with a compatible decapper (e.g., Brooks FluidX Aperio semi-automated decapper or IntelliXcap automated decapper). For SwabFAST (the anterior nares nasal swab collection), we use an oropharyngeal swab with a proximal breakpoint (about 30 mm from the tip of the swab), which allows the swab tip to sit inside the 2 mL cryovial comfortably (Figure 1). For SalivaFAST (saliva collection), we used a funnel or Saliva Collection Aid (Salimetrics), together with the cryovial, to facilitate saliva collection. Some patients might be more comfortable with saliva, while others, due to physical (e.g., lack of saliva), convenience (e.g., forget to avoid eating or drinking before sample collection), or psychological reasons, might prefer nasal swabs. With these two options, patients can choose which type of specimen to use for their COVID-19 tests.

**Figure 1.**
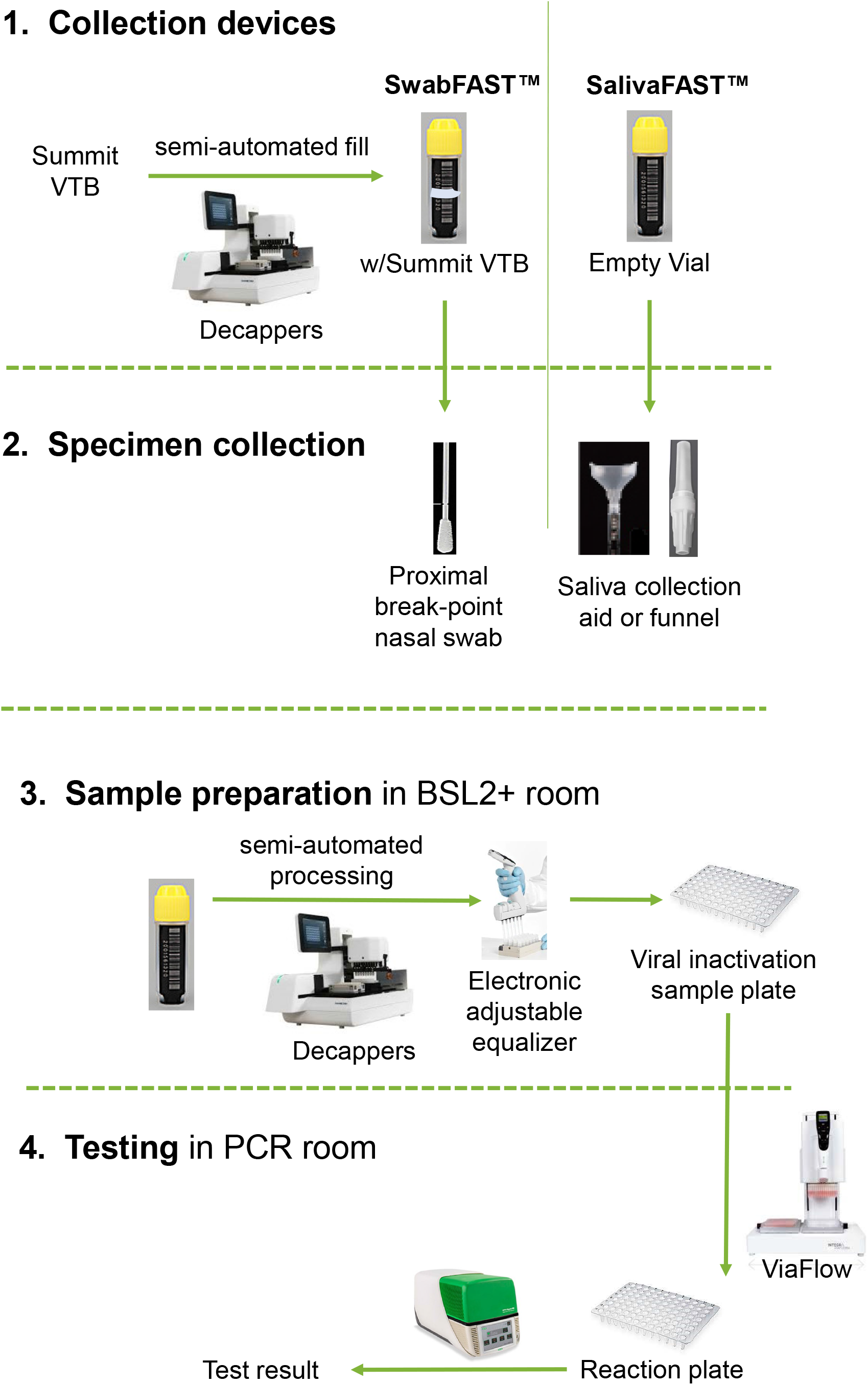
Schematic of SwabFAST and SalivaFAST for high throughput clinical testing. For rapid and high-throughput testing, we use a 2 mL cryovial that has a screw cap with compatible automatic decappers as a sample collection vessel for both anterior nares swab (SwabFAST) samples and saliva (SalivaFAST) samples. For SwabFAST, an oropharyngeal swab with a proximal (about 30 mm) break-point is used for the nasal swab sample collection. After swabbing of patients’ anterior nares, the swab is inserted into the cryovial pre-filled with Summit’s viral transfer buffer (VTB), then broken at the break-point before capping. In SalivaFAST, saliva is collected with either a funnel or a saliva collection aid sitting on top of the cryovial to assist patient with their spit or drool motion. Saliva or swab samples are treated with Summit’s sample prep buffer added at 95°C for 5 minutes in the BSL2+ room. Plates for the PCR machine is setup either manually or with a ViaFlow electronic pipetting machine.

After samples were accessioned, technicians used a decapper and a multi-channel electronic equalizer or a ViaFlow electronic pipetting machine (Integra) to prepare the sample preparation plates in biosafety cabinets in the BSL2+ room (Figure 1, “ BSL2+ room”). Once the sample preparation plates are inactivated, they are moved to the general laboratory for the qPCR testing. Samples in the sample preparation plates will then be transferred to a plate containing PCR master mix with either an equalizer or ViaFlow for RT-qPCR testing (Figure 1, “ PCR room”).

### Optimization of Buffer Component for extraction-free PCR

The central player of extraction-free PCR relies on the efficacy of proteinase K (PK) digestion, which provides a negative selection of breaking down proteins and inactivate DNase and RNases that would otherwise degrade a desired sample of DNA or RNA (*12, 13*). To optimize the best condition for PK activity in either swab or saliva matrix, we have tested a variety of buffer components, commercial swab or saliva collection devices. This is particularly important for swab samples. Unlike saliva, which is able to collect and transport as raw saliva status, swab samples need to be stored in viral transport medium (VTM). However, swab sample in VTM usually requires RNA extraction for SARS-CoV-2 testing.

We thus tested a variety of buffer components, VTM, and a commercial swab collection device-OR100 (DNA Genotek) for extraction-free PCR. Negative swab samples were collected from healthy volunteers and put into each solution. Samples then were spiked into heat-inactivated SARS-CoV-2 virus, mixed with PK according to the SwabFAST protocol in *Material and method* section. As shown in Figure 2A, contrived swab samples in PBS, VTM, or OR100 didn’t generate positive signals at N1 region. Among the positive signals, the contrived swab sample in Tris-Borate-EDTA (TBE) buffer produced the strongest quantification cycle (Cq) value, which comprise the buffer component for our “ viral transport buffer (VTB)”. Similarly, we also tested a variety of buffer components, raw saliva, and a commercial saliva collection device-OM505 (DNA Genotek) for extraction-free PCR. As shown in Figure 2B, contrived saliva samples in OM505 didn’t generate positive signals at N1 region. Among the positive signals, the contrived saliva sample in Tris (2-carboxyethyl) phosphine (TCEP) buffer condition produced the strongest Cq value, which is used to improve PK efficacy in the SalivaFAST protocol.

**Figure 2.**
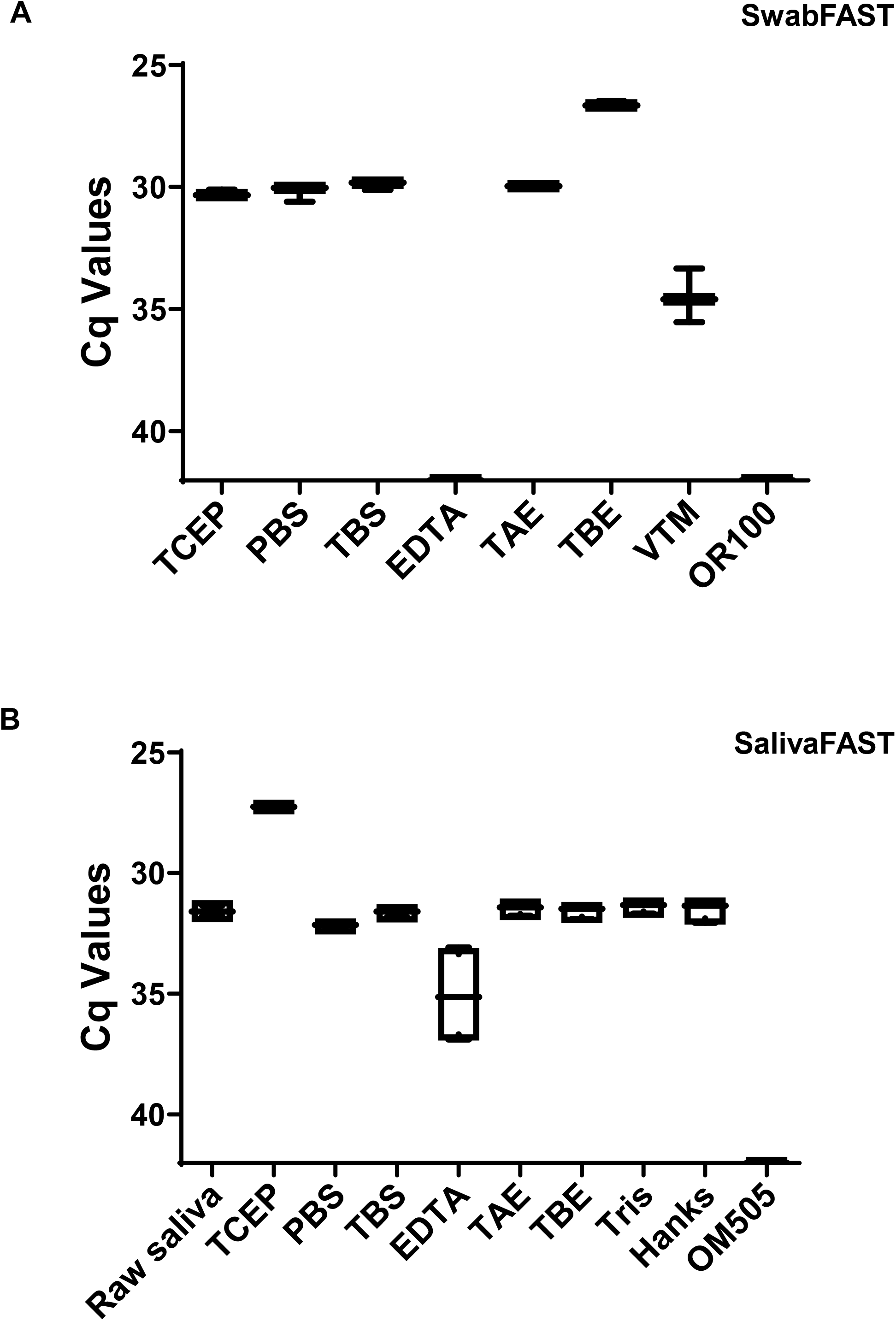
Optimizing buffer components for SwabFAST (A) or SalivaFAST (B). Different buffer components were mixed with 400 cp/μL heat-inactivated SARS-CoV-2 virus contrived swab samples (A) or SARS-CoV-2 contrived saliva samples (B). Cq values at N1 primer/probe were plotted for individual buffer component in swab (A) or saliva (B).

### Stability of VTB

In the SwabFAST test, since the swab collection tubes need to pre-filled VTB before sample collection, there is an issue of VTB stability which may directly affects the quality of the test, assuming that VTB would go through various storage and transportation conditions. To this point, we have tested VTB stability at different temperatures and durations. At higher temperatures, VTB was placed at 4°C, room temperature and 37°C for 32 weeks as of this manuscript, negative swab samples were spiked in heat-inactivated SARS-CoV2 virus at different concentrations, and were tested every four weeks. The result at 32 weeks is shown in Figure 3A. Similarly, at low temperatures, VTB was placed at -80°C, -20°C and 4°C, for 3 weeks as of this manuscript, negative swab samples were spiked in heat-inactivated SARS-CoV2 virus at different concentrations, and were tested every week. The result at 3 weeks is shown in Figure 3B. The Cq value at N1 primer/probe for SARS-CoV-2 contrived samples at different concentrations (4, 40, 400, 4000 copies/µL) remained consistent across all the experimental storage temperatures.

**Figure 3.**
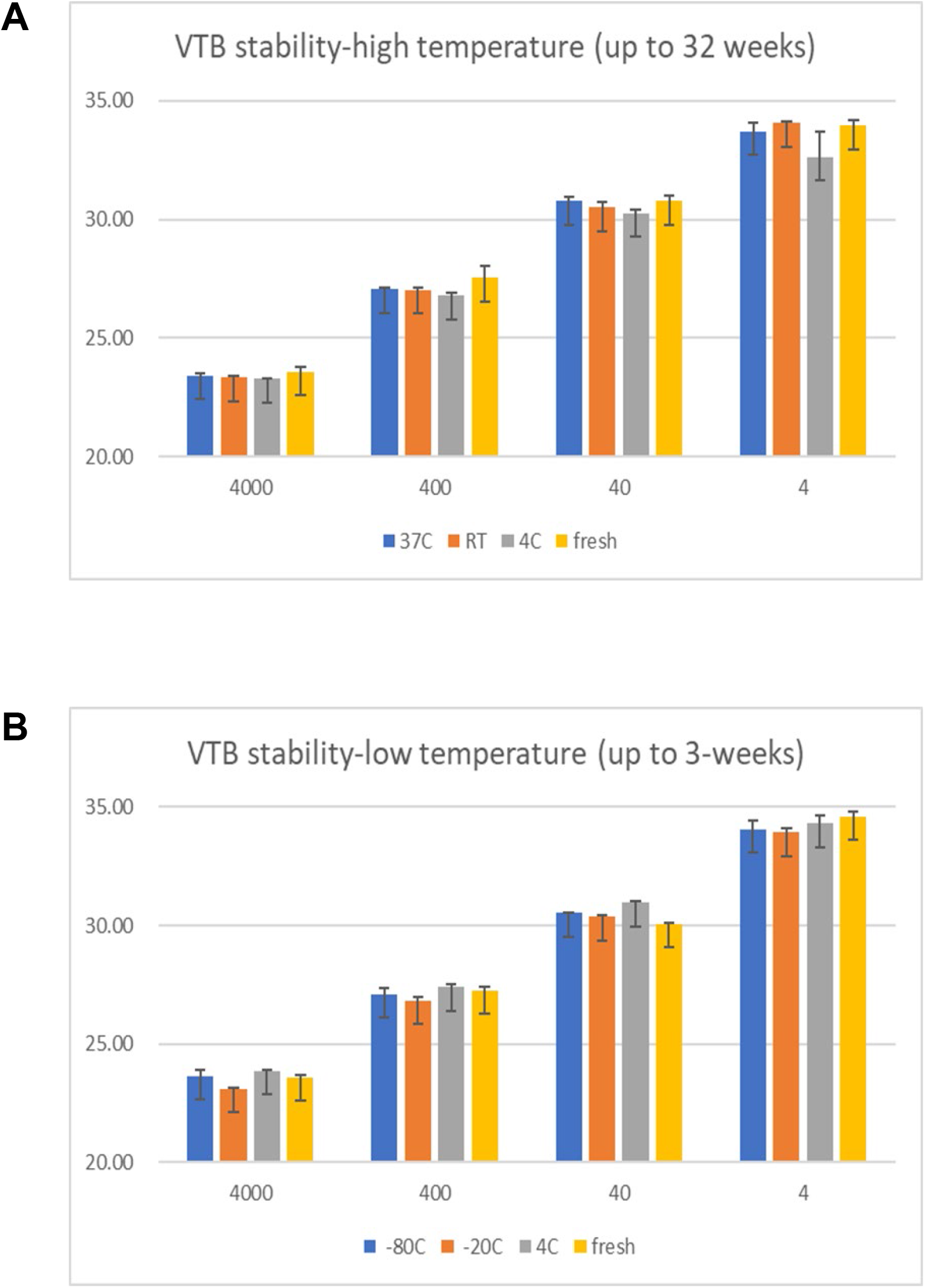
Stability assay of VTB for SwabFAST. VTB was placed at either high temperatures for up to 32 weeks (A), or low temperatures for up to three weeks (B). They were then mixed with SARS-CoV-2 contrived samples at different concentrations (shown on the X-axis). Cq value at N1 primer/probe was used for comparison (shown on the Y-axis).

### Analytical Validation of Extraction-free PCR Testing

To establish analytical validity of the extraction free RT-qPCR assay for nasal swab and saliva specimens, we first compared the results from RT-qPCR testing with or without RNA extraction (Figure 4); we then conducted limit of detection (LoD) studies to determine the lowest detectible concentration of SARS-CoV-2 at which approximately 95% of all true positive samples test positive (Figure 5).

**Figure 4.**
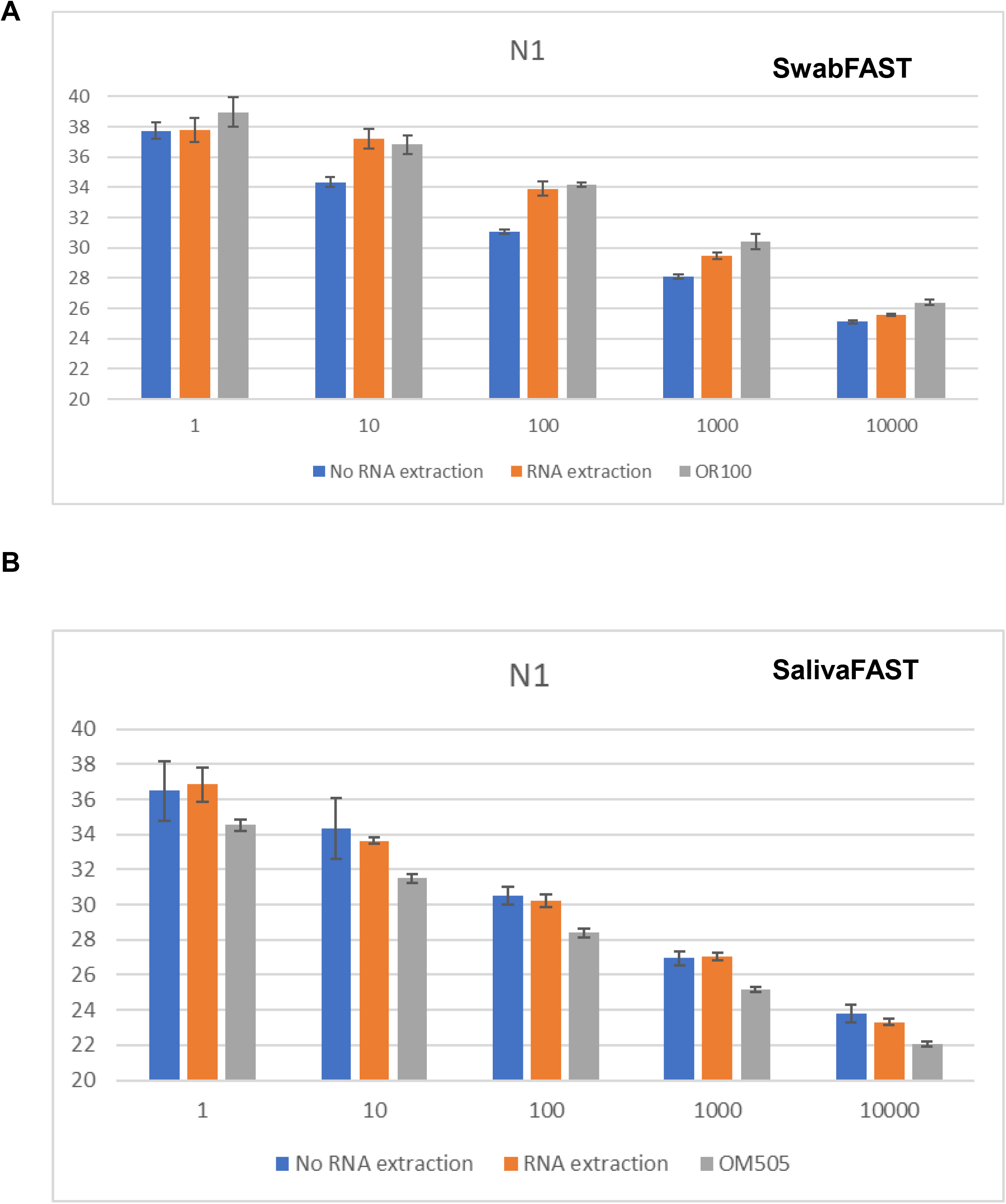
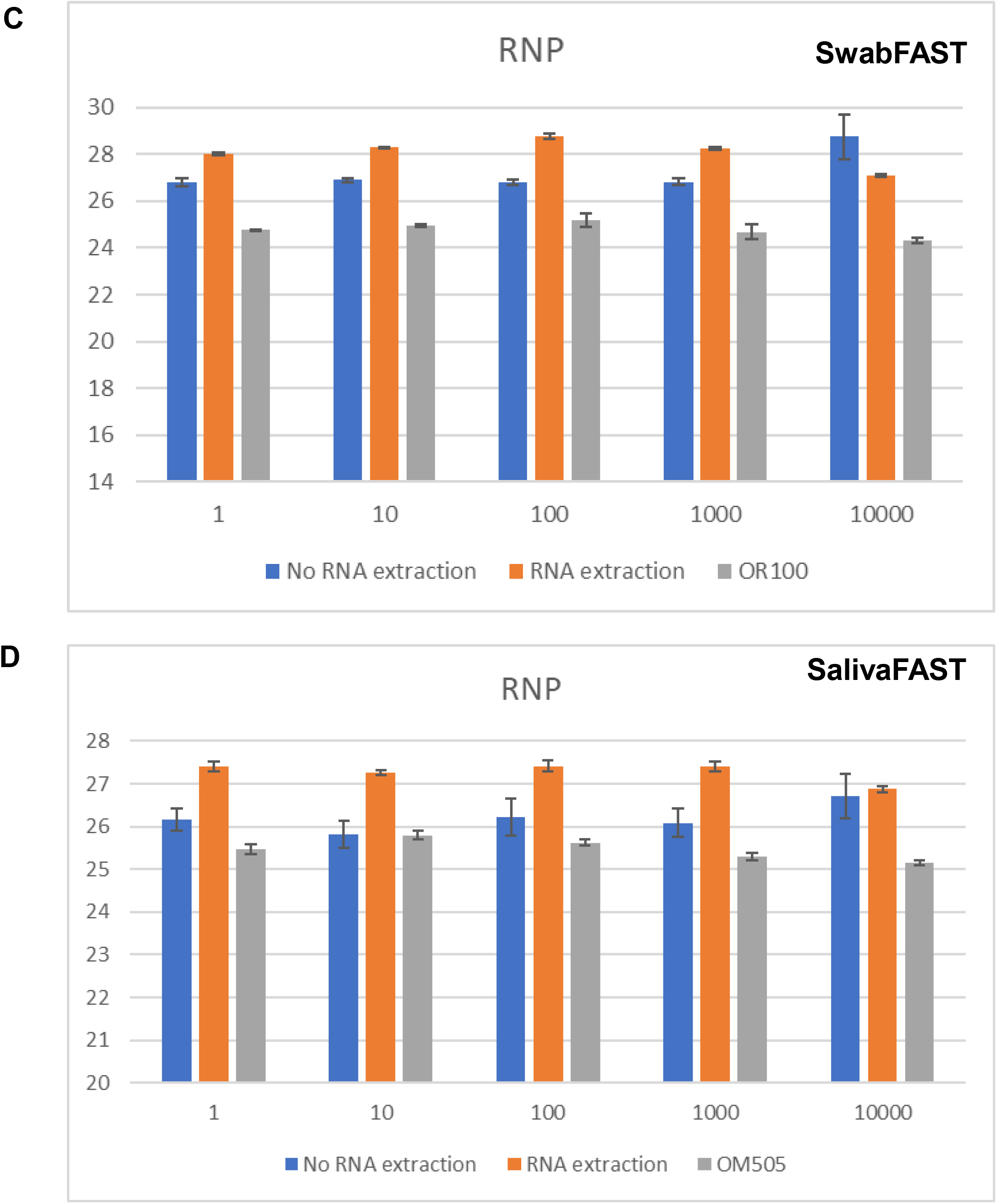
Comparison of test performance with (orange) and without (blue) RNA extraction in contrived samples of swab (A, C) and saliva (B, D). RNA extraction in swab sample (grey) collected in OR100 (A, C), and saliva sample (grey) collected in OM505 (B, D) were also included for comparison. Cq values at N1 region (A, B) or RNP gene (C, D) were plotted in Y-axis.

**Figure 5.**
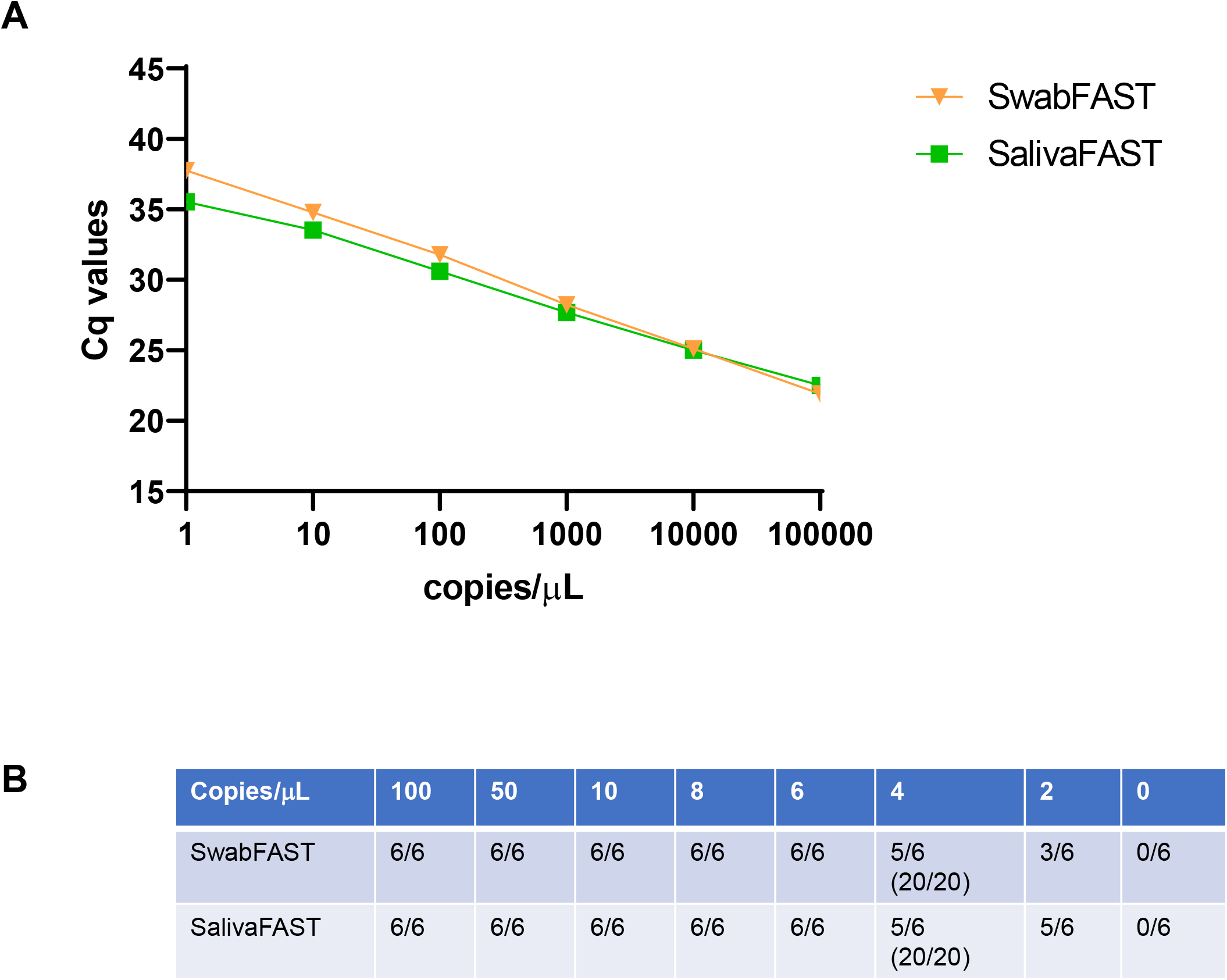
Limit of Detection (LoD) determination. A. Serial dilution curves of SwabFAST (orange) or SalivaFAST (green) were plotted using SARS-CoV-2 contrived samples at concentration ranging from 1 to 100,000 copies/µL. Cq values at N1 primer/probe is shown in Y-axis. B. LoD determination using six replicates of SARS-CoV-2 contrived samples at concentration ranging from 0 to 100 copies/ µL. LoD were further validated in 20 replicates of the contrived samples at concentration of 4 copies/µL.

#### RT-qPCR Results with or without RNA Extraction

To evaluate the extraction-free RT-qPCR methods are equivalent to the RT-qPCR methods with RNA extraction in either swab or saliva samples, we compared the following three methods at different SARS-CoV-2 concentrations (Figure 4): (1) contrived samples (swab or saliva) mixed with buffer analyzed without RNA extraction (blue); (2) contrived samples (swab or saliva) analyzed with RNA extraction (orange); (3) contrived samples (swab or saliva) collected with DNA Genotek’s devices (OR-100 for swab and OM-505 for saliva) analyzed with RNA extraction (gray). The Cq values at the N1 region of SARS-CoV-2 gene (Figure 4A & B) and RNP gene (Figure 4C & D) primer/probe for the three methods at each SARS-CoV-2 concentration (1, 10, 100, 1000, 10000 copies/µL) for both specimen types produce the same qualitative and similar quantitative test results with Cq values across all comparisons.

#### Limit of Detection (LoD)

First, tenfold serially diluted contrived samples at concentration ranging from 1 to 100,000 copies/µL heat-inactivated SARS-CoV-2 viruses were used in independent runs of SwabFAST and SalivaFAST, respectively (Figure 5A). We than further narrow down the range of LoD using six replicates of serially diluted contrived samples at concentration levels of 0, 2, 4, 6, 8, 10, 50, and 100 copies/µL for each specimen type were tested. A preliminary LoD was identified at 4 copies/µL. Confirmation of the LoD was done with 20 replicates at this concentration level. Results show that the LoD of the assays is established at 4 copies/µL, where over 95% of the replicates were tested positive (20/20) for both swab and saliva (Figure 5B).

### Clinical Validation of Extraction-free PCR Testing

#### RT-qPCR Results with or without RNA Extraction

SARS-CoV-2 positive and negative clinical samples were tested with (CDC protocol) (*14*) or without RNA extraction (COVIDFast). 38 SARS-CoV-2 positive and 31 negative swab samples were split in half and the RT-qPCR assay was run with or without RNA extraction. The positive percent agreement (PPV) and the negative percent agreement (NPV) are both 100% (Table 1A). Similarly, 82 SARS-CoV-2 positive and 171 negative saliva samples were split in half and the RT-qPCR assay was run with or without RNA extraction. The PPV and NPV are 98.8% and 99.4% respectively (Table 1B).

**Table 1.** Comparison of test performance with and without RNA extraction in clinical samples. PPA: positive percent agreement; NPA: negative percent agreement.

|  |  | CDC Protocol<br>RNA Extraction* |  |  |
| --- | --- | --- | --- | --- |
|  |  | + | - |  |
| SwabFAST RNA<br>Extraction-free* | + | 38 | 0 | 38 |
|  | - | 0 | 31 | 31 |
|  |  | 38 | 31 | 69 |

PPA = 100% NPA = 100%

B
|  |  | CDC Protocol<br>RNA Extraction* |  |  |
| --- | --- | --- | --- | --- |
|  |  | + | - |  |
| SalivaFAST<br>RNA Extraction-free* | + | 81 | 1 | 82 |
|  | - | 1 | 170 | 171 |
|  |  | 82 | 171 | 253 |

PPA = 98.8 % NPA = 99.4 %

#### Clinical Validation against Another Extraction-free Assay (SalivaDirect)

COVIDFast tests were also clinical validated against SalivaDirect. SwabFAST was validated using a paired SalivaDirect test from the same patient, and SalivaFAST was validated using the same saliva sample collected for SalivaDirect.

179 paired clinical samples—i.e., each testing subject provided one saliva sample and one anterior nares swab sample—from community members were analyzed by SwabFAST in Summit Biolabs and compared to SalivaDirect run by an independent SalivaDirect authorized CLIA lab. The PPV and the NPV are 83.3% and 99.4% respectively (Table 2A). Similarly, 40 raw saliva clinical samples were analyzed by SalivaFAST in Summit Biolabs and compared to SalivaDirect run by an independent SalivaDirect authorized CLIA lab. The PPV and the NPV are 95% and 100% respectively (Table 2B).

**Table 2.** Clinical validation of SwabFAST (A) and SalivaFAST (B) against SalivaDirect. PPA: positive percent agreement; NPA: negative percent agreement.

|  |  | SalivaDirect |  |
| --- | --- | --- | --- |
|  |  | Positive | Negative |
| SwabFAST | Positive | 5 | 1 |
|  | Negative | 1 | 178 |
|  |  | 6 | 179 |
|  | PPA | 83.3% (5/6) |  |
|  | NPA | 99.4% (178/179) |  |

B
|  |  | SalivaDirect |  |
| --- | --- | --- | --- |
|  |  | Positive | Negative |
| SalivaFAST | Positive | 19 | 0 |
|  | Negative | 1 | 20 |
|  |  | 20 | 20 |
|  | PPA | 95% (19/20) |  |
|  | NPA | 100% (20/20) |  |

### Real-world Experience of Cumulative Clinical Testing Results for COVIDFast

Since launch COVIDFast as an LDT, we have performed a total of 409,883 tests by December 23, 2021. Unexpectedly, the percentage of SwabFAST (83.62%, 342,734/409,883) being performed is much higher than that of SalivaFAST (16.38%, 67,149/409,883). The total positive rate of COVIDFast is 8.10%, with 7.71% positive in SwabFAST and 10.09% positive in SalivaFAST. (Table 3)

**Table 3.** Summary of clinical testing results for COVIDFast as laboratory development tests (LDTs) in Summit Biolabs as of December 28, 2021.

|  | SwabFAST | SalivaFAST | COVIDFast |
| --- | --- | --- | --- |
| SARS-Cov2 positives/Total tested | 26,425/342,734 (7.71%) | 6,775/67,149 (10.09%) | 33,200/409,883 (8.10%) |

## DISCUSSION

In this paper, we describe the COVIDFast assay, which includes SwabFAST and SalivaFAST, for the rapid detection of the viral infection of SARS-CoV-2. We reported the high throughput setup, buffers for extraction-free PCR, analytical and clinical validation findings and real world experience of COVID testing in Colorado. The extraction step in conventional PCR creates a major bottle neck in the diagnostic process, as it remains both manually laborious and expensive, and further increases the chances of accidental contamination and human error. Bypassing the RNA extraction step offers the advantage of curtailing additional resources normally spent on industrial RNA extraction kits, human capital involved in the extraction steps, and all aspects of lab operations in support of this step. Therefore, the RNA extraction-free method of detecting SARS-CoV-2 is a prime candidate for building a rapid and high-throughput testing system.

One critical piece in the COVIDFast assay is the buffer, which made extraction-free RT-qPCR testing for SARS-CoV-2 possible. The buffer composition is specific to the type of sample, i.e. buffers for SwabFAST or for SalivaFAST. Furthermore, the buffer composition for SwabFAST also serves as a transport buffer, such that the swab is immediately placed within collection vessel containing the buffer, hence the name “ viral transfer buffer” or “ VTB.” Although we are not the first to report extraction-free PCR testing for SARS-CoV-2 using saliva as a specimen (*14-18*), to our knowledge, we are the first to create a direct-to-PCR SARS-CoV-2 assay for nasal swabs. We demonstrated the efficacy of the buffer for both specimen types, making COVIDFast very flexible and versatile. In a clinical context where the majority of individuals still prefer a shallow nasal swab (anterior nares) due to reasons such as its ease of sample collection (i.e., no requirements of food and beverage restriction prior to testing) and ready availability (i.e., some patients have trouble generating saliva), having choices for sample types translates into more testing and better detection of SARS-CoV-2 in the pandemic. Further, we demonstrated that the buffer is stable over time in varying conditions, which bears significance for clinical testing as sample integrity during transportation is a critical step in ensuring testing quality. Being able to use the buffer as a transport medium is extremely meaningful in a central lab testing model that is still dominant in the U.S. and many other countries.

We also assembled a high throughput system containing specific types of collection vessels, devices, and lab equipment, all compatible with the extraction-free assay we designed. This high throughput system, when coupled with the extraction-free feature of the assays, drastically improves on the speed at which RT-qPCR tests can be resulted, making the “ rapid” in “ rapid PCR testing” a reality. For example, COVIDFast tests have a 24-hour turnaround time after sample receipt at Summit’s CLIA lab at a volume of >5,000/day. Specifically, to our knowledge, we are the first to use a small cryovial with an externally threaded screw cap as a sample collection vessel. The adoption of a standard cryovial not only makes liquid handling automation possible, for example, by using tube-compatible equipment such as automated decappers, it also made sample storage easier by eliminating the sample transfer step. The screw cap helps prevent contamination. In addition, using the same cryovials for both swab and saliva sample types simplify the accessioning and sample preparation steps immensely, creating additional efficiency in the system. As part of the collection kits, a funnel or saliva collection aid is used to facilitate saliva collection (SalivaFAST), and a low-breakpoint (approx. 30 mm from the tip of the swab) nylon oropharyngeal swab is used (SwabFAST), which allows the swab to be inserted into the tube after use. The compact sizes of the collection vessels and devices make transportation by mail or courier much easier. In short, the high throughput system contributes to the rapid PCR testing promise made to the patients and their doctors.

In combination, the extraction-free feature of the assays and the high throughput system made the rapid direct-to-PCR testing affordable. With the RNA-extraction step gone, the automation of the sample handling and preparation lead to both cost savings in materials and man power, which can be passed onto the patients and the entire public health system. An affordable and accurate test means that patients can be tested more frequently, thus catching infections earlier and preventing further spread of the virus (*1, 4*). Last, COVIDFast tests are the first successful clinical application of the RNA extraction-free RT qPCR diagnostics platform. Future applications and expansion of the nucleic acid-extraction free platform with a similar high-volume system can include other viruses, such as influenzas, and may expand to bacteria and fungus detection.

## MATERIALS AND METHODS

### Source of Samples

Human samples in this study is approved by the Salus IRB #Summit-COVID-SLV-1.

#### Contrived samples

All the contrived samples described in this paper were assembled by spiking the heat-inactivated SARS-Cov-2 virus (VR-1986HK, ATCC) into known negative anterior nares nasal swab or saliva samples.

#### Samples for clinical validation with SalivaDirect

Samples comparing SwabFAST (anterior nares nasal swab) and SalivaDirect (raw saliva) came from an IRB-approved clinical site for SARS-CoV-2 testing. Patients were students, faculty, staff, and community members of a university. Patients were instructed and supervised by trained healthcare workers during the collection of samples. Anterior nasal swabs were collected with an oropharyngeal swab, which was then broken and tip inserted into a 2 mL cryogenic vial filled with 1 ml of VTB, and paired saliva was collected according to the SalivaDirect protocol (*13*). All samples were transported to the labs at ambient temperature within 24 hours and analyzed within 36 hours of collection in the laboratory. Swab samples were analyzed according to SwabFAST protocol detailed below at Summit Biolabs, and saliva samples were analyzed by a SalivaDirect authorized CLIA lab.

Samples comparing SalivaFAST and SalivaDirect came from a CLIA lab authorized to perform SalivaDirect tests on clinical samples. Remnants of 40 clinical saliva samples that were tested positive or negative for SARS-CoV-2 were assigned a blind random number (1-40) and sent to the Summit Biolabs for testing with SalivaFAST using the protocol detailed below.

#### COVIDFast Clinical Sample Collection

The healthcare worker ensures all patient information, including name, date of birth, and additional information required by state reporting rules, is filled out properly and tied to the collection vessel (2 mL cryovial) and the biohazard bag that the collection vessel will be stored before processed by the laboratory.

##### Anterior nares swab sample collection for SwabFAST

The anterior nares nasal swab was collected under the supervision of a trained healthcare worker. Before collection, patients were given a quick review of the collection procedure such as one recommended by the FDA (https://tinyurl.com/nasalswab1-2). An oral or nares swab with a proximal (30 mm) break point was used to swab 10 times on each nare and breaks the swab inside the tube at the proximal breakpoint close to the swab. The 2 mL cryovial was filled with 1 mL of VTB and has an externally threaded screw cap, which has pre-printed barcodes for identification use.

##### Saliva sample collection for SwabFAST

Saliva samples were collected from individuals by having them spit into a 2 mL cryovial. A saliva collection aide (Salimetrix) or support funnel were used in tandem with the cryovial.

The healthcare worker replaces the cap of the tube and make sure it’s securely fastened. The sample were then be placed in a sealed individual biohazard bag under room temperature before being transported to the lab.

### RNA extraction

Total RNA was extracted from either swab or saliva using the QIAamp Viral RNA extraction kit (Qiagen)

### Extraction-free SARS-CoV-2 RT-qPCR Diagnostics

#### Sample preparation

Once samples (in 2 mL cryovials) passed the accessioning step (e.g., patient information is complete, no leakage, no dry swabs), and sample information entered into the lab information management system (LIMS), they were placed on a platform rocker in hold position at 60 rpm until a lab technical from the sample preparation team fetches the samples.

Sample preparation entails aliquoting the sample into a 96 plate pre-filled with either 1) a mix of saliva preparation buffer and PK (Promega) for saliva samples, or 2) PK (Promega) alone for swab samples. For SalivaFAST, 30 µL from a single saliva sample is mixed with 5 µL saliva preparation buffer and 5 µL PK in each well of the plate (total 40 µL per well). For SwabFAST, 35 µL from a single swab sample is mixed with 5 µL proteinase K per well. The difference here is that swab samples are collected and transported in the buffer, thus, no further buffer is needed.

The prepared sample mix plate is then placed on a digital microplate shaker at 500 RPM for one minute, then on a thermal cycler at 95°C for five minutes for heat-inactivation (*19*). It may then be placed on hold in a 4°C refrigerator for further processing.

#### Duplex PCR reagent preparation and reaction

The CDC 2019 Novel Coronavirus (2019-nCoV) RT-qPCR assay (*14*) was implemented and adapted for extraction-free SARS-CoV-2 testing (COVIDFast). We duplexed N1 region and ribonuclease P (RNP) primer/probes in one assay. The PCR master mix consists of 10 μL Luna Universal Probe One-Step Reaction Mix (NEB), 1 μL Luna Warmstart RT enzyme Mix (NEB), and 1.5 μL of N1/RNP primers/probes (6.7 uM for primers and 1.7 uM FAM-labeled N1 probe and 1.7 uM ATTO-647 labeled RNP probe, IDT). An aliquot of 7.5 μL pre-treated sample (saliva or swab) from the sample preparation step was added into each well. Same amount of positive control (IDT synthetic 2019-SARS-CoV-N control, 4000 copies/uL), negative control (IDT Hs-RPP30 control,4000 copies/μL) for SARS-CoV-2, and no-template control (NTC—water) were included in each plate. The PCR plate was run on the CFX Opus 96 machine (Biorad) using the following thermal profile: Step 1: 55°C 10 minutes, 1 cycle; Step 2: 95°C 1 minute, 1 cycle; and Step 3: 95°C 10 sec then go to 60°C 30 sec (+ plate read at both FAM channel for N1 target & Cy5 channel for RNP target) for 40 cycles.

#### Data interpretation

Interpretation of the Cq values for N1 and RNP targets is based on a Cq value threshold of 36 for N1 and 35 for RNP, as derived from the clinical validation studies for COVIDFast tests:

|  | Cq: N1 (FAM channel) | Cq: RNP (Cy5 channel) |
| --- | --- | --- |
| Detected (SARS-CoV-2 Positive) | $\leq 36$ | Any number or NaN (Not a Number) |
| Not Detected (SARS-CoV-2 Negative) | $> 36$ | $\leq 35$ |
| Invalid | $> 36$ | $> 35$ |

## Data Availability

A master transfer agreement (MTA) was not used to access samples in this study. All data associated with this study are available in the main text or the supplementary materials.

## Acknowledgments

The authors would like to thank CU Innovations, COVIDCheck Colorado, Colorado Mesa University, and Warrior Diagnostics. The authors would also like to thank the advice from David Brunel, Pauline Gee, and Stanley Lapidus.

## Funding

Funding of the study is provided by University of Colorado Anschutz Medical Campus SPARK program, University of Colorado Anschutz Medical Campus Chancellor Discovery Innovation Fund (CDI Fund), Colorado Office of Economic Development and International Trade (OEDIT) Advanced Industry Award, and Summit Biolabs, Inc.

## Author contributions

Conceptualization (YQ, BB, XY, JZ, BH, SL)

Investigation (YQ, LL, AH, RT, SY, FG, AL, XY, SL)

Methodology (YQ, LL, BH, XY, SL)

Data Curation (YQ, LL AH, RT, SY, FG, AL, XY, SL)

Formal Analysis (DG, BH, SL) Writing – original draft (XY)

Writing – review & editing (XY, BH, SL)

Resources (SL)

Funding acquisition (SL)

## Competing interests

DG, BB, XY, JZ, BH, and SL own equity in Summit Biolabs, Inc.

## Notes

### Author Declarations

Salus IRB approval #Summit-COVID19-SLV-1

